# Reduction of Microbial Diversity During Artisanal Fermentation and Detection of Multidrug-Resistant *Salmonella* in Commercially Imported Beef in Puerto Rico

**DOI:** 10.64898/2026.06.01.26354615

**Authors:** Milliani Carmona-Ortiz, Joyce Cartagena-Teruel, Ariadna Maldonado-Maldonado, Lorenid Martínez-Pacheco, Lizel Ortiz-Cartagena, Gian Rodríguez-Plata, Gustavo Santiago-Collazo

**Affiliations:** Department of Biology, University of Puerto Rico, Ponce, PR 00716

**Keywords:** fermented meat products, embutidos, *Salmonella*, *Enterobacter*, antimicrobial resistance, multidrug resistance, food safety, Puerto Rico, artisanal production

## Abstract

Artisanally produced *embutidos* are a culturally significant fermented meat product widely consumed in Puerto Rico, yet their microbiological safety remains largely uncharacterized. This preliminary study evaluated the effect of artisanal fermentation on microbial diversity and assessed the presence of potentially pathogenic and antimicrobial-resistant *Proteobacteria* in locally produced embutidos. Raw pork (locally sourced) and beef (commercially imported) were obtained from retail supermarkets and processed at a small-scale production facility under standard artisanal conditions. Surface sampling using RODAC contact plates on TSA, MAC, and SDA media was performed before and after fermentation. Fermentation reduced overall microbial diversity in both meat types. Two Gram-negative isolates recovered from pre-fermentation samples were characterized using selective and differential media (MAC, MSA, EMB) and the IMViC biochemical test series. A *Salmonella* species was presumptively identified from imported beef, and an *Enterobacter* species from locally sourced pork. Kirby-Bauer disk diffusion testing revealed that the *Salmonella* isolate was resistant to five antibiotics (ampicillin, methicillin, penicillin, streptomycin, tetracycline) and showed intermediate susceptibility to chloramphenicol and gentamicin. The *Enterobacter* isolate was resistant to five antibiotics (ampicillin, methicillin, penicillin, streptomycin, tetracycline) and showed intermediate susceptibility to chloramphenicol and gentamicin. Both isolates met MDR criteria, highlighting the need for enhanced microbiological oversight of artisanally produced embutidos in Puerto Rico.

## Introduction

Fermented meat products, commonly known as embutidos, represent a cornerstone of Puerto Rican culinary culture and are consumed year-round across the island. Unlike industrially produced products, the majority of embutidos available in Puerto Rico are produced artisanally by small-scale producers who often source raw meat directly from local supermarkets, operating outside the scope of rigorous microbiological quality control. As a result, the microbiological safety of these products remains largely unknown, posing a potential public health concern that has received limited scientific attention in the local context.

Fermentation has long been recognized as one of the most effective traditional preservation strategies for meat products, contributing to extended shelf life, enhanced organoleptic properties, and the reduction of spoilage and pathogenic microorganisms through acidification and microbial competition (Wang et al., 2021; Leistner, 2000). However, the effectiveness of fermentation as a barrier against pathogenic bacteria is not absolute, particularly when the raw material harbors microorganisms capable of tolerating or adapting to the fermentation environment. Understanding whether artisanal fermentation practices are sufficient to reduce microbial diversity and control pathogenic genera is therefore essential for evaluating the safety of locally produced embutidos.

Of particular concern is the presence of pathogenic *Proteobacteria*, such as *Salmonella* spp. and *Enterobacter* spp., which have been associated with raw meat and meat products globally (Terentjeva et al., 2017; Conceição et al., 2023). Beyond their pathogenic potential, these genera are increasingly documented as carriers of antimicrobial resistance determinants, raising serious public health concerns within the One Health framework (WHO, 2015; CDC, 2022). In the United States, the USDA Food Safety and Inspection Service (FSIS) actively monitors *Salmonella* in meat and poultry through the Pathogen Reduction/Hazard Analysis and Critical Control Point (PR/HACCP) framework (USDA-FSIS, 2013). The National Antimicrobial Resistance Monitoring System (NARMS), a collaborative initiative between USDA, FDA, and CDC, tracks antimicrobial resistance in foodborne enteric bacteria from retail meats and food-producing animals (NARMS, 2024). Despite these federal surveillance mechanisms, artisanally produced embutidos in Puerto Rico falls largely outside the scope of formal microbiological oversight, representing an important gap in the FSIS regulatory framework.

Despite the cultural and economic relevance of artisanally produced embutidos in Puerto Rico, no preliminary microbiological characterization of these products from raw material to fermented product has been reported using locally sourced pork and imported beef. This knowledge gap limits evidence-based risk assessment and the development of targeted safety interventions for small-scale producers. The present study aimed to conduct a preliminary microbiological evaluation of artisanally fermented embutidos produced from local pork and imported beef.

Specifically, we sought to: (1) assess whether fermentation reduces overall microbial diversity by comparing microbiological profiles before and after fermentation; (2) isolate and presumptively identify potentially pathogenic *Proteobacteria* from pre-fermentation samples using differential and selective media (MAC, MSA, EMB) and the IMViC test series; and (3) determine the antimicrobial resistance profiles of the identified isolates through antibiogram testing. Findings provide preliminary evidence relevant to public health risk assessment for artisanally produced fermented meat products in Puerto Rico.

## Materials and Methods

### Sample Collection and Source

Raw meat was purchased directly from local supermarkets in Puerto Rico, reflecting the actual sourcing practices of artisanal embutido producers. Two meat types were obtained: locally sourced pork (*Sus scrofa domesticus*) and commercially imported beef. Samples were transported under refrigeration (≤ 4°C) to a local small-scale embutido producer, where the fermentation process was carried out under standard artisanal production conditions. All sampling was performed at the production facility immediately before and after the fermentation process. All laboratory procedures were carried out by undergraduate students enrolled in the Undergraduate Research Course BIOL 3108 at the University of Puerto Rico at Ponce, under the direct supervision of the course instructor and Principal Investigator.

### Experimental Design

A before-and-after fermentation design was employed to assess changes in the microbiological profile associated with the fermentation process. Pre-fermentation samples (raw meat) served as the baseline for microbial load assessment and pathogen isolation. Post-fermentation samples (finished fermented product) were used to evaluate whether the fermentation process resulted in a measurable reduction of overall microbial diversity.

### Surface Sampling Using RODAC Plates

Surface microbial sampling was performed using Replicate Organism Detection and Counting (RODAC) contact plates directly on the meat surface at the production facility. Although RODAC plates are primarily designed for surface sampling of environmental and food contact surfaces, this method was employed as a practical preliminary approach to capture the microbial communities present on the meat surface at the point of production.

Three types of RODAC plates were used: Tryptic Soy Agar (TSA): For enumeration of total aerobic mesophilic bacteria. MacConkey Agar (MAC): Selective and differential medium for Gram-negative enteric bacteria. Sabouraud Dextrose Agar (SDA): For detection of yeasts and molds. Fungal diversity was detected but was not further characterized in the present study, as fungal ecology falls outside the scope of this report.

TSA and MAC plates were incubated at 37°C for 24–48 hours. SDA plates were incubated at 25°C for 48–72 hours. Colony counts and morphologies were recorded for all plates. It is acknowledged that RODAC contact plates represent a surface sampling approach and may not fully capture the microbial communities present within the meat matrix; results should therefore be interpreted as preliminary and indicative of the surface microbiome at the time of sampling.

### Isolation of Gram-Negative Bacteria

Following incubation, MacConkey agar RODAC plates were used as the primary source for the isolation of Gram-negative bacteria. Morphologically distinct colonies were selected from MAC plates and sub-cultured onto fresh MacConkey agar plates to enrich for Gram-negative diversity and obtain pure isolates. Two colonies presenting distinct and contrasting morphologies one from the pre-fermentation pork sample and one from the pre-fermentation beef sample were selected for further biochemical characterization based on their differential reactions on MAC agar. Each isolate was subsequently sub-cultured onto Tryptic Soy Agar to confirm purity prior to the biochemical test series.

### Presumptive Biochemical Identification

Each pure isolate was subjected to a series of differential and selective tests to presumptively identify the bacterial genus and species.

### Selective and Differential Media

Each isolate was independently streaked onto three selective and differential media to assess biochemical and growth characteristics. MacConkey Agar (MAC) was used for Gram-negative selection and lactose fermentation differentiation, with pink or red colonies indicating lactose fermentation and colorless colonies indicating lactose-negative reactions. Mannitol Salt Agar (MSA) was included as a selective medium for *Staphylococcus* spp., allowing confirmation of the absence of halotolerant Gram-positive contaminants among the selected isolates. Eosin Methylene Blue Agar (EMB) was used for Gram-negative differentiation; production of a metallic green sheen is characteristic of *Escherichia coli*, while other colony morphologies and colorations on EMB support the presumptive identification of alternative enteric genera.

### IMViC Biochemical Test Series

The IMViC test series was performed to differentiate among members of the family *Enterobacteriaceae* based on four core metabolic reactions. The Indole test was conducted using tryptone broth with Kovács reagent; a red ring at the broth surface indicates a positive result, reflecting the ability of the organism to produce indole from tryptophan. The Methyl Red (MR) test was performed using MR-VP broth with Methyl Red indicator; a red color development (pH ≤4.4) constitutes a positive result, indicative of stable acid end-products from mixed acid fermentation. The Voges-Proskauer (VP) test was carried out in MR-VP broth using Barritt’s reagents A and B; a red-pink color reaction indicates acetoin production, a characteristic feature of organisms that use the butanediol fermentation pathway. The Citrate utilization test was performed on Simmons Citrate Agar; blue coloration of the medium or visible growth indicates a positive result, reflecting the organism’s ability to use citrate as its sole carbon source. Results were interpreted collectively using standard *Enterobacteriaceae* identification keys to assign presumptive genus-level identification to each isolate.

### Antimicrobial Susceptibility Testing

Antimicrobial resistance profiles of both isolates were determined using the disk diffusion method (Kirby-Bauer), in accordance with the guidelines of the Clinical and Laboratory Standards Institute (CLSI). Bacterial suspensions were prepared to a ∼0.5 McFarland turbidity standard and uniformly inoculated onto Mueller-Hinton Agar (MHA) plates by sterile swabbing. Antibiotic disks were positioned on inoculated plates, which were subsequently incubated at 37°C for 18–24 hours. Inhibition zone diameters were measured and interpreted as Susceptible (S), Intermediate (I), or Resistant (R) according to current CLSI breakpoints. All susceptibility tests were performed in triplicate. Mean zone diameters and standard deviations are reported. A panel of seven (7) antibiotics representing clinically and veterinary relevant antimicrobial classes was evaluated. Ampicillin (10mcg), Chloramphenicol (30mcg), Gentamycin (10mcg), Penicillin (10mcg), Streptomycin (10mcg) and, Tetracycline (30mcg). Isolates demonstrating resistance to antibiotics belonging to three or more antimicrobial classes were classified as multidrug-resistant (MDR), consistent with the standard definition proposed by Magiorakos et al. (2012).

## Results

### Effect of Fermentation on Microbial Diversity

Comparison of RODAC contact plates obtained before and after fermentation revealed a notable shift in the microbiological profile of both meat types. Pre-fermentation plates displayed a heterogeneous colonial morphology, characterized by the presence of multiple colony types differing in size, color, and surface texture, indicative of a diverse microbial community (Figure 1A). Post-fermentation plates, in contrast, exhibited a markedly homogeneous colonial appearance, with predominance of a single or limited colony morphotype, suggesting a substantial reduction in overall microbial diversity associated with the fermentation process (Figure 1B). This pattern was observed consistently across both locally sourced pork and commercially imported beef samples.

**Figure 1.**
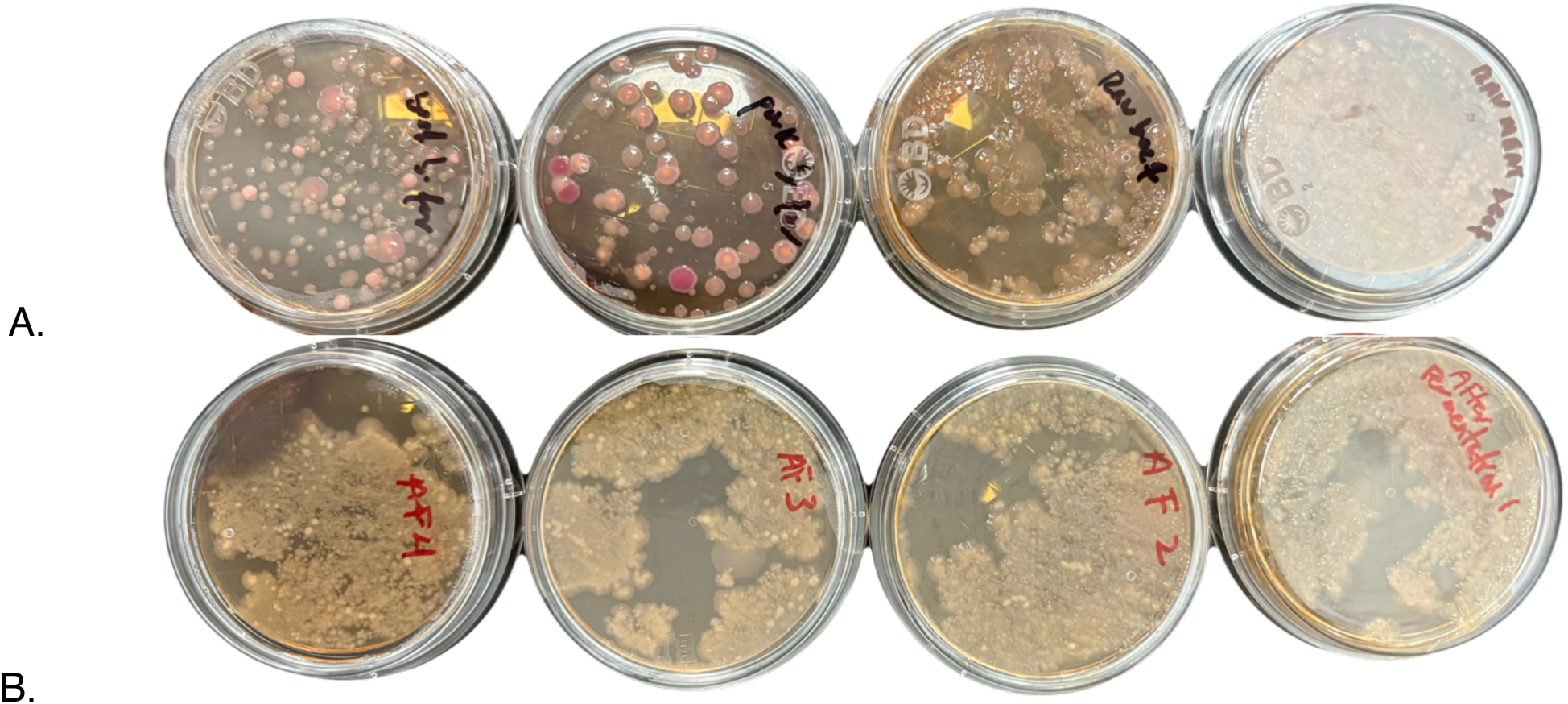
Effect of artisanal fermentation on microbial diversity as assessed by RODAC contact plate sampling. Representative RODAC plates on Tryptic Soy Agar (TSA) obtained from (A) locally sourced pork and commercially imported beef pre-fermentation (B) locally sourced pork and commercially imported beef post-fermentation. Pre-fermentation plates display heterogeneous colonial morphology indicative of diverse microbial communities. Post-fermentation plates exhibit homogeneous colonial morphology consistent with a reduction in overall microbial diversity. Plates were incubated at 37°C for 24–48 hours.

### Isolation and Colonial Morphology of Target Isolates

Initial surface sampling was performed using TSA RODAC plates (Figure 1); morphologically distinct colonies were subsequently sub-cultured onto MacConkey agar for Gram-negative isolation and differential characterization (Figure 2). The isolate recovered from imported beef (Unknown 1) produced pink colonies on MAC agar, consistent with lactose fermentation (Figure 2A). The isolate recovered from locally sourced pork (Unknown 2) also produced pink colonies on MAC agar; however, the plate exhibited a characteristic orange discoloration, a pattern associated with acid overproduction from lactose fermentation by members of the genus *Enterobacter* (Winn et al., 2006) (Figure 2B). On Eosin Methylene Blue agar, Unknown 1 produced dark/black colonies (Figure 2C) and Unknown 2 produced pink colonies (Figure 2D), consistent with differential enteric reactions on EMB and supporting distinct genus-level identities for the two isolates (Forbes et al., 2007).

**Figure 2.**
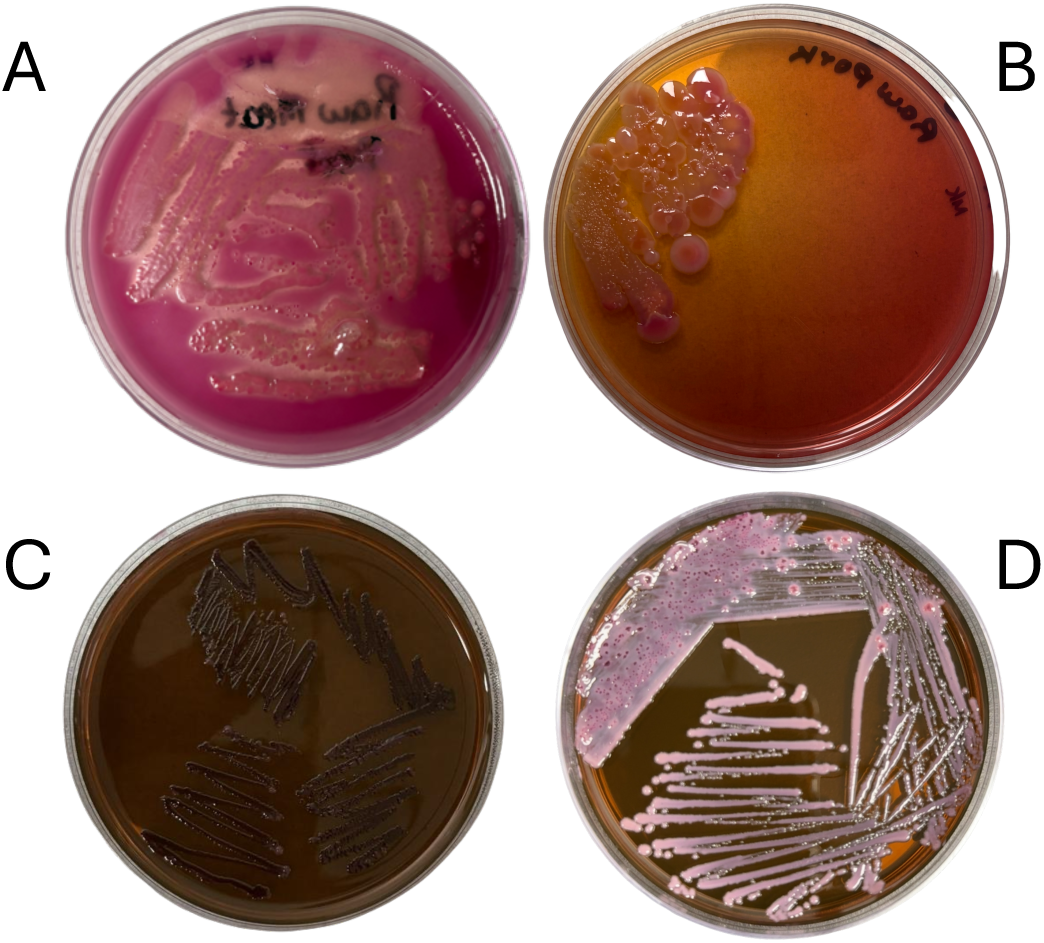
Colonial morphology of Gram-negative isolates on MacConkey Agar (MAC) and Eosin Methylene Blue Agar (EMB). (A) Unknown 1 (beef isolate) on MAC, displaying positive lactose fermentation. (B) Unknown 2 (pork isolate) on MAC, displaying partially positive lactose reaction with characteristic orange agar discoloration consistent with variable lactose fermentation in members of the genus *Enterobacter* (Winn et al., 2006). (C) Unknown 1 on EMB, displaying non-*E. coli* colonial morphology without metallic green sheen. (D) Unknown 2 on EMB, displaying non-*E. coli* colonial morphology without metallic green sheen. Plates were incubated at 37°C for 24–48 hours.

### Biochemical Characterization

#### Unknown 1: Presumptive Salmonella spp. (Beef Isolate)

SIM medium results revealed negative indole production, positive motility, and positive hydrogen sulfide (H₂S) production, evidenced by blackening of the medium. Triple Sugar Iron (TSI) agar results were positive for butt acidification with gas production and formation of a black precipitate, consistent with H₂S production (Winn et al., 2006). The Methyl Red (MR) test yielded a partial positive result and the Voges-Proskauer (VP) test was negative. Nitrate reduction was positive. The combined biochemical profile H₂S production, gas formation in TSI, negative indole, positive motility, and partial MR reaction is consistent with the presumptive identification of a *Salmonella* species (Table 1B) (Whitman, 2015; Forbes et al., 2007).

**Table 1A.** Results of selective and differential media testing for Gram-negative isolates recovered from pre-fermentation meat samples. Selective and differential medium reactions of Unknown 1 (commercially imported beef) and Unknown 2 (locally sourced pork). MacConkey Agar (MAC) was used for Gram-negative selection and lactose fermentation differentiation. Mannitol Salt Agar (MSA) was used to assess halotolerance and mannitol fermentation, confirming the absence of *Staphylococcus* spp. among the selected isolates. Eosin Methylene Blue Agar (EMB) was used for Gram-negative differentiation; absence of metallic green sheen excludes *Escherichia coli* and supports the presumptive identification of alternative enteric genera. Unknown 1 exhibited limited growth on MSA without mannitol fermentation, suggesting incidental salt tolerance rather than the characteristic halotolerance of *Staphylococcus* spp. (+) = positive reaction; (−) = negative reaction. All plates were incubated at 37°C for 24–48 hours.

| Test / Medium | Unknown 1 (Beef) | Unknown 2 (Pork) |
| --- | --- | --- |
| **MacConkey (MAC)** | Positive (+); lactose fermenter | Partially positive; variable lactose reaction with orange agar discoloration |
| **MSA — Growth in NaCl** | Positive (limited growth; no mannitol fermentation) | Negative |
| **MSA — Mannitol Fermentation** | Negative | Negative |
| **EMB** | Negative metallic sheen; non-*E. coli* morphology | Negative metallic sheen; non-*E. coli* morphology |

**Table 1B.** IMViC biochemical reactions and confirmatory test results for Unknown Sample #1 (raw beef) and Unknown Sample #2 (raw pork). Indole production was assessed using Kovács reagent. Methyl Red (MR) and Voges-Proskauer (VP) reactions were performed using MR-VP broth with their respective indicators; VP was negative for both isolates, consistent with the absence of acetoin production. Citrate utilization was assessed on Simmons Citrate Agar; both isolates were positive. H₂S production and motility were assessed simultaneously using SIM medium. Triple Sugar Iron (TSI) agar reactions are reported as slant/butt: K = alkaline (−); A = acid (+). Presumptive genus-level identification was assigned based on the combined biochemical profile per standard *Enterobacteriaceae* identification keys (Winn et al., 2006; Forbes et al., 2007; Whitman, 2015). (+) = positive; (−) = negative; (+/−) = partially positive.

| Test | Reagent / Medium | Unknown 1 (Beef) | Unknown 2 (Pork) |
| --- | --- | --- | --- |
| **Indole (I)** | Tryptone broth + Kovács reagent | Negative (–) | Negative (–) |
| **Methyl Red (M)** | MR-VP broth + MR indicator | Partially positive (+/–) | Negative (–) |
| **Voges-Proskauer (V)** | MR-VP broth + Barritt's A & B | Negative (–) | Negative (–) |
| **Citrate (C)** | Simmons Citrate Agar | Positive (+) | Positive (+) |
| **H<sub>2</sub>S Production** | SIM medium | Positive (+) | Negative (–) |
| **Motility** | SIM medium | Positive (+) | Positive (+) |
| **TSI – Slant/Butt** | Triple Sugar Iron Agar | K (–) / A (+) | K (–) / A (+) |
| **TSI – Glucose** | Triple Sugar Iron Agar | Positive (+) | Positive (+) |
| **TSI – Sucrose** | Triple Sugar Iron Agar | Negative (–) | Negative (–) |
| **TSI – Lactose** | Triple Sugar Iron Agar | Negative (–) | Negative (–) |
| **TSI – H<sub>2</sub>S** | Triple Sugar Iron Agar | Positive (+) | Negative (–) |
| **Presumptive ID** |  | *Salmonella* spp. | *Enterobacter* spp. |

#### Unknown 2: Presumptive Enterobacter spp. (Pork Isolate)

SIM medium results revealed negative indole production, negative H₂S production, and positive motility. TSI agar results were positive for butt acidification without gas production or black precipitate formation, confirming absence of H₂S activity. The Methyl Red (MR) and Voges-Proskauer (VP) tests were both negative. Nitrate reduction was positive. The biochemical profile absence of H₂S and gas in TSI, negative indole, negative MR and VP, positive motility, and characteristic orange discoloration on MacConkey agar is consistent with the presumptive identification of an *Enterobacter* species (Table 1B) (Whitman, 2015; Winn et al., 2006).

### Antimicrobial Susceptibility

Kirby-Bauer disk diffusion testing was performed in accordance with CLSI guidelines (CLSI, 2023), demonstrating distinct antimicrobial resistance profiles between the two isolates. The presumptive *Salmonella* isolate (Unknown 1, beef) was resistant (R) to five of the seven antibiotics tested ampicillin, methicillin, penicillin, streptomycin, and tetracycline and intermediate (I) for chloramphenicol (mean 13.72 mm) and gentamicin (mean 15.08 mm) (Figure 3, Table 2). Resistance spanning three antimicrobial classes (beta-lactams: ampicillin, methicillin, penicillin; aminoglycosides: streptomycin; tetracyclines: tetracycline) confirmed classification of this isolate as multidrug-resistant (MDR) per Magiorakos et al. (2012). The presumptive *Enterobacter* isolate (Unknown 2, pork) demonstrated to be resistant (R) to ampicillin, methicillin, penicillin, streptomycin, and tetracycline; intermediate for chloramphenicol (mean 14.26 mm) and gentamicin (mean 15.41 mm), also meeting MDR criteria. (Figure 3, Table 2)

**Figure 3.**
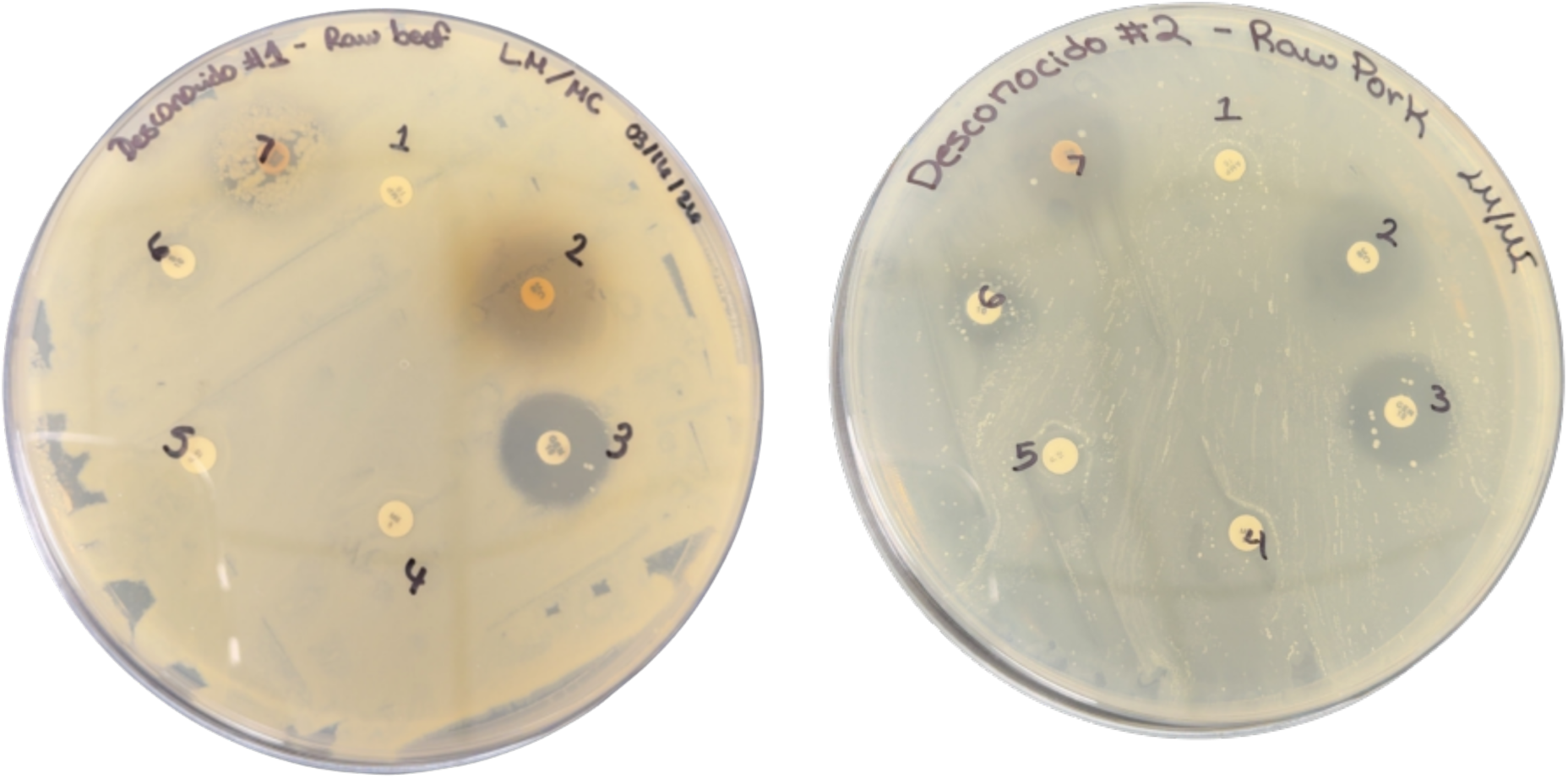
Kirby-Bauer disk diffusion antibiogram results for both isolates on Mueller Hinton Agar (MH). (A) Unknown 1 (presumptive *Salmonella*, beef isolate) showing resistance to ampicillin (1), methicillin (4), penicillin (5), streptomycin (6), and tetracycline (7) (absent or reduced inhibition zones) and susceptibility to chloramphenicol and gentamicin (clear inhibition zones). (B) Unknown 2 (presumptive *Enterobacter*, pork isolate) showing resistance to beta-lactam antibiotics ampicillin (1), methicillin (4), penicillin (5); and streptomycin (6) and tetracycline (7) and intermediate to chloramphenicol (2) and gentamicin (3). Plates were incubated at 37°C for 18–24 hours. Zone diameters were interpreted per CLSI (2023) breakpoints.

**Table 2.** Antimicrobial susceptibility profiles of Unknown 1 (presumptive *Salmonella* spp., beef isolate) and Unknown 2 (presumptive *Enterobacter* spp., pork isolate) determined by Kirby-Bauer disk diffusion on Mueller-Hinton Agar. Zone diameters are reported in millimeters (mm). Interpretations were assigned as Susceptible (S), Intermediate (I), or Resistant (R) per Clinical and Laboratory Standards Institute breakpoints (CLSI, 2023). MDR classification was assigned to isolates demonstrating resistance to antibiotics belonging to three or more distinct antimicrobial classes per Magiorakos et al. (2012). Conc. = concentration; R = resistant; S = susceptible; I = intermediate; MDR = multidrug-resistant. *R* = Resistant, no CLSI breakpoints established for Enterobacteriaceae.* All tests were performed in triplicate. Values represent mean zone diameters ± standard deviation (SD).

| Antibiotic | Disk | U1 Mean ± SD (mm) | U1 Interp. | U2 Mean ± SD (mm) | U2 Interp. |
| --- | --- | --- | --- | --- | --- |
| Ampicillin | 10 μg | 0.00 ± 0.00 | R | 0.00 ± 0.00 | R |
| Chloramphenicol | 30 μg | 13.72 ± 5.45 | I | 14.26 ± 3.40 | I |
| Gentamicin | 10 μg | 15.08 ± 5.00 | I | 15.41 ± 2.50 | I |
| Methicillin | 5 μg | 0.00 ± 0.00 | R* | 0.00 ± 0.00 | R* |
| Penicillin | 10 μg | 0.00 ± 0.00 | R* | 0.00 ± 0.00 | R* |
| Streptomycin | 10 μg | 0.00 ± 0.00 | R | 5.54 ± 4.85 | R |
| Tetracycline | 30 μg | 0.00 ± 0.00 | R | 10.41 ± 4.30 | R |
| **MDR Classification** |  |  | **MDR ✓** |  | **MDR ✓** |

## Discussion

### Fermentation as a Microbial Diversity Reduction Strategy

The observed shift from heterogeneous to homogeneous colonial morphology following fermentation is consistent with the well-documented role of lactic acid fermentation as a selective preservation mechanism in meat products (Wang et al., 2021; Leistner, 2000). Acidification of the meat matrix suppresses diverse microbial communities while favoring acid-tolerant organisms, typically lactic acid bacteria. These findings support the premise that artisanal fermentation, even under non-standardized conditions, can effectively reduce overall microbial diversity. However, the persistence of potentially pathogenic isolates in pre-fermentation samples underscores that fermentation alone cannot be relied upon as the sole food safety intervention when the raw material harbors resistant organisms. Biochemical characterization of the Gram-negative isolates recovered from pre-fermentation samples further revealed notable discrepancies between selective and confirmatory media, warranting careful interpretation of individual test results.

### Biochemical Identification and Profile Interpretation

The biochemical profile of Unknown 1 is consistent with the genus *Salmonella*, as characterized in standard microbiological references (Whitman, 2015; Winn et al., 2006). H₂S production in SIM and TSI reflects thiosulfate reductase activity, a hallmark of *Salmonella* (Forbes et al., 2007). Gas in TSI, negative indole, partial MR, and negative VP further support this identification. Detection of presumptive *Salmonella* in commercially imported beef suggests contamination introduced through the import supply chain (Terentjeva et al., 2017), consistent with NARMS retail beef surveillance data (NARMS, 2024). The biochemical profile of Unknown 2 aligns with the genus *Enterobacter*, particularly based on characteristic orange discoloration on MAC resulting from strong lactose-fermenting acid-overproducing activity typical of this genus (Winn et al., 2006; Jorgensen et al., 2015). Negative H₂S, negative indole, and positive motility are consistent with established *Enterobacter* keys (Whitman, 2015; Forbes et al., 2007). The negative VP result, while atypical, does not exclude this genus, as VP-negative strains have been documented (Jorgensen et al., 2015), reinforcing the need for molecular confirmation.

### Antimicrobial Resistance Profiles and Public Health Implications

The MDR profile of the presumptive *Salmonella* isolate, resistant to beta-lactam antibiotics, streptomycin, and tetracycline, with intermediate susceptibility to chloramphenicol and gentamicin, is consistent with MDR patterns previously documented in *Salmonella* recovered from food animal products globally (Terentjeva et al., 2017; Conceição et al., 2023) and aligns with antimicrobial resistance trends reported by NARMS in retail meat surveillance across the United States (NARMS, 2024). FSIS has identified *Salmonella* control as a top food safety priority; however, artisanally produced products in U.S. territories are not systematically included in federal sampling programs (USDA-FSIS, 2013), representing a critical surveillance gap. The global threat of antimicrobial resistance in foodborne pathogens further underscores the public health significance of this finding (WHO, 2015; CDC, 2022) and highlights the urgent need to extend federal surveillance frameworks to small-scale artisanal producers in Puerto Rico.

The presumptive *Enterobacter* isolate also met MDR criteria, demonstrating resistance to beta-lactam antibiotics, streptomycin, and tetracycline, with intermediate susceptibility to chloramphenicol and gentamicin, a profile with significant clinical implications. Intermediate susceptibility to chloramphenicol and gentamicin in both isolates warrants clinical caution, as intermediate results may reflect reduced drug efficacy at standard dosing and could mask the emergence of full resistance under sustained therapeutic pressure (CLSI, 2023). *Enterobacter* species are recognized opportunistic pathogens, particularly in immunocompromised individuals, and their detection in artisanally processed meat products warrants close attention within the One Health framework (Jorgensen et al., 2015). The breadth of resistance observed in both isolates suggests prior selective pressure from antimicrobial use within the production chain, potentially operating at multiple points across both import and local supply networks (Ahmed et al., 2025).

Notably, both isolates exhibited largely overlapping MDR profiles despite originating from distinct meat sources, commercially imported beef and locally sourced pork, respectively. This convergence suggests that antimicrobial selective pressure may not be restricted to imported products but may also be present within the local pork supply chain in Puerto Rico. These findings underscore the need for systematic microbiological surveillance of artisanally produced embutidos regardless of meat origin and support the integration of small-scale artisanal producers into existing federal food safety monitoring frameworks.

### Limitations and Future Directions

Several limitations merit consideration. First, RODAC contact plates are suited for surface monitoring and may underestimate the full microbial diversity within the meat matrix (Jorgensen et al., 2015). Second, species-level identification was based exclusively on biochemical characterization; molecular confirmation via 16S rRNA sequencing and targeted PCR will be employed (Whitman, 2015). Third, the sample size one production batch per meat type restricts generalizability. Future studies should expand sampling to multiple producers, batches, and seasons to characterize the broader microbiological landscape of artisanally produced embutidos in Puerto Rico. Notably, considerable variability was observed among triplicates for certain antibiotic-isolate combinations (e.g., chloramphenicol for Unknown 1: 10.16, 11.00, 20.00 mm; SD = 5.45), which may reflect technical variation inherent to surface-sampled colonies or inconsistencies in inoculum density. Future work should include standardized inoculum preparation and replicate confirmation to reduce measurement variability. Integration with NARMS data and alignment with USDA-FSIS frameworks would contextualize local findings within national surveillance systems (NARMS, 2024; USDA-FSIS, 2013). Metagenomic approaches would additionally allow comprehensive resistome and microbial diversity profiling before and after fermentation

## Data Availability

All data produced in the present work are contained in the manuscript

## Abbreviations

TSA: Tryptic Soy Agar
MAC: MacConkey
SDA: Sabouraud Dextrose Agar
RODAC: Replicate Organism Detection and Counting
IMViC: Indole, Methyl Red
Voges: Proskauer, Citrate
CLSI: Clinical and Laboratory Standards Institute
MDR: Multi Drug Resistant

## Acknowledgements.

This study was conducted as part of an undergraduate research course at the University of Puerto Rico at Ponce (UPRP) under the supervision and leadership of Dr. Gustavo Santiago-Collazo who served as Principal Investigator. The authors thank the participating students for their contributions to sample collection, laboratory work, and data recording. The authors also thank Charcutería Coameña for letting us their facilities for sampling and fermentation process and drying process.

